# Acceptability of COVID-19 vaccination among health care workers in Ghana

**DOI:** 10.1101/2021.03.11.21253374

**Authors:** Martin Wiredu Agyekum, Grace Frempong Afrifa-Anane, Frank Kyei-Arthur, Bright Addo

**Author notes:** **Corresponding Author** Bright Addo, Department of Sociology and Social Work, Kwame Nkrumah University of Science and Technology, Kumasi-Ghana. /.

## Abstract

The acceptance or otherwise of the COVID-19 vaccine by health care workers can influence the uptake of COVID-19 vaccines among the general population as they are a reliable source of health information. In this study, we sought to determine the acceptability of COVID-19 vaccines among health care workers in Ghana. Using a cross-sectional design, we collected data from 234 health care workers through a self-administered online survey from 16 January to 15 February 2021. Descriptive, bivariate and multivariate analyses using binary logistic regression were performed using STATA version 15. The results showed that 39.3% of health care workers had the intention of receiving the COVID-19 vaccine. Factors such as sex, category of health care workers, relative being diagnosed with COVID-19, and trust in the accuracy of the measures taken by the government in the fight against COVID-19 proved to be significant predictors of the acceptability of the COVID-19 vaccine. Concerns about the safety of vaccines and the adverse side effects of the vaccine were identified as the main reasons why health care workers would decline uptake of the COVID-19 vaccine in Ghana. The self-reported low intention of health care workers to accept the COVID-19 vaccine in Ghana requires the urgent call of the Government of Ghana and other stakeholders to critically address health care workers’ concerns about the safety and adverse side effects of COVID-19 vaccines, as this would increase vaccine uptake. Interventions must also take into consideration sex and the category of health care workers to achieve the desired results.

## 1. Introduction

In March 2020, the novel coronavirus disease (COVID-19) was declared a global pandemic after its emergence in Wuhan, China in November 2019. Globally, as of 22 February 2021, there were 111,102,016 confirmed cases of COVID-19 and 2,462,911 deaths [1]. In Ghana, the total number of confirmed cases as of 21 February was 80,253 with 577 deaths [2]. Ghanaian health care workers have not been spared the brunt of the pandemic. Since the emergence of COVID-19 in Ghana on 12 March 2020, approximately 1,629 nurses and midwives have been infected with the virus and 4 have died as of 14 February 2021 [3]. Also, more than 450 doctors and dentists have been infected with COVID-19 and 7 have died [4].

Governments globally have imposed several measures and protocols to help halt the spread of COVID-19 such as travel bans, the mandatory wearing of nose marks, lockdowns, social distance, and frequent washing of hands with soap and water, among others. To further halt the spread and deaths associated with COVID-19, researchers and pharmaceutical companies are collaborating to develop safe and effective vaccines for mass vaccination. Vaccination is an effective way of combating infectious conditions [5]. As of 12 February 2021, 66 vaccines were in the clinical development stage while 176 were at the pre-clinical development stage [6]. Globally, five vaccines have been deemed safe and effective for human use as of 12 February 2021 namely: Pfizer, Oxford/AstraZeneca, Moderna, Janssen, and Sputnik V [7-9].

Due to the inadequate supply of COVID-19 vaccines globally, governments have prioritised high-risk groups to receive the initial supply of vaccines. These high-risk groups include health care workers, older persons especially those with chronic co-morbid conditions, and those in essential services [10]. Health care workers are at a high risk of contracting the COVID-19 disease due to their direct or indirect contact with bodily secretions, COVID-19 patients/clients, visitors, and other health care workers who may have been exposed to COVID-19 [11, 12].

Nobody is more familiar with the impact of COVID-19 than health care workers who are at the forefront of the battle against the disease. Nonetheless, many health care workers continue to indicate refusal or hesitancy in the uptake of the COVID-19 vaccine. The availability of COVID-19 vaccines therefore may not translate into its uptake. Although governments will provide the vaccines, their uptake is voluntary [7]. Indeed, several studies have demonstrated that not all health care workers are ready to accept COVID-19 vaccines when made available in their country [13-15]. For example, a Hong Kong study found that about 40% of nurses had the intention to accept the COVID-19 vaccine if available [16]. Similarly, a multi-country study in France, Belgium, and Canada examining attitudes of health workers towards COVID-19 vaccination found that approximately 40% of health workers in Belgium (Wallonia and Brussels) were willing to vaccinate themselves if COVID-19 vaccines were available [15].

In sub-Saharan Africa (SSA), a study conducted in the Democratic Republic of Congo found that approximately 28% of health care workers were willing to receive the COVID-19 vaccine if available [17]. Differences in the decision to accept COVID-19 vaccines when made available have also been noted in these studies. Compared to female health care workers, male health care workers are more likely to accept COVID-19 vaccines. Also, compared to nurses and midwives, medical doctors were found to be more likely to accept the COVID-19 vaccine if made available [13, 14, 17, 18]. Reasons for hesitancy to accept COVID-19 vaccines have been identified to include concerns over vaccine safety and side effects, speed of vaccine development/approval, [16, 19, 20].

The Government of Ghana (GoG) has publicly articulated its decision to initiate the procurement of the Oxford/AstraZeneca and Sputnik V COVID-19 vaccines for use in the country. Health care workers have been identified as the first recipients of the vaccine. Since the announcement was made for the purchase of COVID-19 vaccines in Ghana, there has been a mixed response as to whether or not to accept or refuse vaccination by several people in the country, including health care workers. Surprisingly, limited studies have been conducted in Ghana to determine the extent to which health care workers will accept vaccination.

Health care workers are a reliable source of information on vaccination to patients [13, 21], therefore their acceptance or otherwise of the COVID-19 vaccine may influence the uptake of COVID-19 among the general population. Against this backdrop, this study seeks to assess and identify the determinants of COVID-19 acceptability among health care workers in Ghana. As Ghana expects to receive her first batch of COVID-19 vaccines in March 2021, examining the acceptance of COVID-19 vaccination among health workers would help researchers and policymakers to design appropriate interventions to reduce vaccine hesitancy among health workers and the general population.

## 2. Methods

### 2.1 Study setting

Ghana is a lower-middle-income country and it has an estimated 30.9 million population. It is bounded by Burkina Faso on the North, the Gulf of Guinea on the South, Togo on the East, and Cote d’Ivoire on the West. Ghana has 16 administrative regions. As of 2017, Ghana had 5,421 Community-based Health Planning and Services (CHPS), 998 clinics, 140 district hospitals, 1004 health centres, and 357 hospitals. Health facilities and health workers in Ghana are disproportionately found in the Ashanti and Greater Accra regions, especially in urban areas. For instance, out of the 357 hospitals in Ghana, 128 (35.9%) are found in the Ashanti region and 99 (27.7%) in the Greater Accra region [22]. Also, Ghana has 4,016 doctors as of 2017 and almost two-fifth (39.4%) are found in the Greater Accra region while one-fifth (20.5%) are found in the Ashanti region. In terms of critical health staff (such as community health nurses, enrolled nurses, medical officers, and pharmacist, among others), Ghana has 90,703 personnel and 13,438 (14.8%) and 13,120 (14.5%) respectively are found in Ashanti and Greater Accra regions [22].

### 2.2 Study Design

This survey is a cross-sectional study conducted among health care workers in Ghana using both convenient and snowballing sampling techniques. Google Form was used to design an online self-administered questionnaire and it was disseminated through WhatsApp to health care workers’ platforms. In addition, a snowball sampling technique was used to reach out to health workers by encouraging respondents to forward or share the online survey link to other health workers. This approach was adopted because of the existing nature of the pandemic, as it offers appropriate social distancing and excludes movements of researchers or participants [23]. To be included in the study, the individual must be a health professional working in a registered health care setting and must consent and be willing to participate in the survey. Participation in the study was voluntary and anonymity was assured. The data were collected from 16 January 2021 to 15 February 2021.

### 2.3 Measurement and Variables

Acceptance in this study was defined as the intention to accept to take the COVID-19 vaccine. This was the dependent variable and it was assessed using the question: “if an approved COVID-19 vaccine becomes available, would you take it yourself?” and the responses are “Yes” and “No”.

The questionnaire developed covered four sections: (i) socio-demographic characteristics, (ii) COVID-19 experience, (iii) trust in the fight against COVID-19 pandemic, and (iv) intention to vaccinate. The socio-demographic characteristics included measures such as age (20-29 years; 30-39 years; 40 and above), sex (Males and Female), religious affiliation (Christians; non-Christians), highest level of schooling (Certificate; Diploma; First degree; Post-graduate) category of health care workers (nurses and midwives; Allied health workers; Medical doctors), marital status (Never married; Married), subjective household wealth (More money than you need; Just enough money; Less money than you need), received any vaccination apart from childhood immunisation (Yes, No), and chronic illness status (Yes; No).

Regarding COVID-19 experience, respondents were asked about their previous contact with a COVID-19 patient, whether a member of their household, relatives, friends or neighbours has been diagnosed with COVID-19. Respondents were also asked about their COVID-19 status and their perceived risk of contracting the COVID-19 disease. The response for each question was Yes and No.

To measure social trust in the fight against COVID-19 pandemic, nine questions were used and assessed using a 5-point Likert scale (strongly disagree, disagree, neutral, agree and strongly agree). The questions were: I trust in the authorities in the fight against COVID-19; I trust in the information provided by media in the fight against COVID-19; I trust in our health system and hospitals in the fight against COVID-19; I trust in the accuracy of the measures taken by the government in the fight against COVID-19; I trust in the correct implementation of the measures in the fight against COVID-19; I trust in the appropriateness of the economic measures taken with respect to COVID-19; I think that we are more successful than the Western countries in the fight against COVID-19; I think that COVID-19 pandemic will lose its effect with summer; COVID-19 has showed that the countries deemed to be powerful are not that powerful).

These questions have been validated for the measure in previous studies [24]. However, some of the questions were modified to reflect our local context. Confirmatory factor analysis was performed to evaluate the reliability validity of the Scale. A Cronbach’s alpha coefficient was used to check for the reliability of the social trust scale. The Cronbach alpha coefficient of the scale was 0.8675. A Cronbach alpha coefficient higher than 0.60 indicates high reliability [24].

### 2.4 Statistical Analysis

The data were analysed using STATA version 15. Descriptive statistics were used to describe the socio-demographic characteristics of health workers. Also, Pearson’s chi-square was used to determine the association between intention to vaccinate and socio-demographic characteristics, COVID-19 experience, and social trust in the fight against the COVID-19 pandemic. The dependent variable was dichotomous so multivariate binary logistic regression analysis was used to identify the determinants of intention to vaccinate. All variables were considered statistically significant at 95% confidence interval (*p < 0.05*).

## 3. Results

### 3.1 Sample characteristics

There were a total of 234 health care workers who completed the online survey. Of the 234 health care workers, more than half (56%) were aged 30-39 years and the least proportion (11.1%) were aged 40 years and above. The majority (63.2%) of the health care workers were females and a higher proportion of females (66.9%) significantly indicated non-acceptance of COVID-19 vaccine compared to males (50%; p < 0.05). The majority of the health care workers were Christians and about seven out of ten (76.5%) were living in an urban area. Regarding educational attainment, the highest proportion (36.8%) of health care workers had a bachelor’s degree, about one-fifth (20.5%) had a postgraduate degree, and 14.1% had a certificate. Also, the majority (64.5%) of health care workers were nurses and midwives, 23.5% were allied health care workers and 12% were medical doctors. In terms of marital status, more than half (56.8%) of the health care workers were married, while 43.2% were not married. A higher proportion of those who were never married (69.3%) indicated non-acceptance of the COVID-19 vaccine compared to those who were married (54.1%; p < 0.05). About four-fifth (79.1%) of the health care workers had received vaccination in their lifetime and 90.6% of the health care workers reported no chronic disease (Table 1).

**Table 1.** Background characteristics of health care workers

|  | n (%) | Acceptance of COVID-19 Vaccine |  | *P*-value |
| --- | --- | --- | --- | --- |
|  |  | No<br>n (%) | Yes<br>n (%) |  |
| Age |  |  |  |  |
| 20-29 | 77 (32.9) | 49 (63.6%) | 28 (36.4%) | 0.265 |
| 30-39 | 131 (56.0) | 81 (61.8%) | 50 (38.2%) |  |
| 40 and above | 26 (11.1) | 12 (11.5%) | 92 (88.5%) |  |
| Sex |  |  |  |  |
| Male | 86 (36.8) | 43 (50.0%) | 43 (50.0%) | 0.011** |
| Female | 148 (63.2) | 99 (66.9%) | 49 (33.1%) |  |
| Religion |  |  |  |  |
| Christians | 216 (92.3) | 129 (59.7%) | 87 (40.3%) | 0.297 |
| Non-Christians | 18 (7.7) | 13 (72.2%) | 5 (27.8%) |  |
| Educational level |  |  |  |  |
| Certificate | 33 (14.1) | 20 (60.6%) | 13 (39.4%) | 0.054** |
| Diploma | 67 (28.6) | 45 (67.2%) | 22 (32.8%) |  |
| First degree | 86 (36.8) | 56 (65.1%) | 30 (34.9%) |  |
| Postgraduate | 48 (20.5) | 21 (43.8%) | 27 (56.2%) |  |
| Category of health care workers |  |  |  |  |
| Nurse and midwife | 151 (64.5) | 98 (64.9%) | 53 (35.1%) | 0.039** |
| Allied health workers | 55 (23.5) | 33 (60.0%) | 22 (40.0%) |  |
| Medical doctors | 28 (12.0) | 11 (39.3%) | 17 (60.7%) |  |
| Marital status |  |  |  |  |
| Never Married | 101 (43.2) | 70 (69.3%) | 31 (30.7%) | 0.019** |
| Married | 133 (56.8) | 72 (54.1%) | 61 (45.9%) |  |
| Place of residence |  |  |  |  |
| Urban | 179 (76.5) | 104 (58.1%) | 75 (41.9%) | 0.144 |
| Rural | 55 (23.5) | 38 (69.1%) | 17 (30.9%) |  |
| Received any vaccination apart from childhood immunisation |  |  |  |  |
| Yes | 185 (79.1) | 112 (60.5%) | 73 (39.5%) | 0.931 |
| No | 49 (20.9) | 30 (61.2%) | 19 (38.8%) |  |
| Any chronic illness |  |  |  |  |
| Yes | 22 (9.4) | 11 (50.0%) | 11 (50.0%) | 0.281 |
| No | 212 (90.6) | 131 (61.8%) | 81 (38.2%) |  |
| $p < .1^*, p < .05^{**}, p < .01^{***}$ | | | | |

### 3.2 Acceptance of COVID-19 vaccine

Out of the 234 health care workers who participated in the study, about two-fifth (n=92, 39.3%) of them indicated acceptance of COVID-19 vaccine if available while more than half (n=142, 60.7%) indicated non-acceptance of COVID-19 vaccine if available.

### 3.3 Health care workers experience with COVID-19

About 60.7% of the respondents indicated that they had not been in contact with any COVID-19 patient whiles 39.2% had been in contact with a COVID-19 patient. A higher proportion of those who had not been in contact with a COVID-19 patient (66.5%) indicated no acceptance of the COVID-19 vaccine compared to those who had contacted a patient (52.2%; *p < 0.05*). Also, eight out of ten (83.8%) respondents indicated that no member in their households had been diagnosed with COVID-19. A higher proportion of those whose household member(s) had been diagnosed with COVID-19 (57.9%) indicated acceptance of the COVID-19 vaccine (*p < 0.05*). In addition, 68.8% of the respondents had none of their relatives been diagnosed with COVID-19, 19.2% had their relatives been diagnosed with COVID-19 and 12% of them were uncertain if their relatives had been diagnosed with COVID-19. Also, the highest proportion of health care workers (44.4%) indicated that their friends had been diagnosed with COVID-19. Approximately 48% of the respondents had been tested for COVID-19; seven out of ten (70.8%) tested negative, while about one-tenth (9.7%) did not receive their test results. Also, 47.9 % of the respondents indicated a great chance of getting COVID-19 while less than one-tenth (6.4%) indicated no risk at all. The majority of the study respondents (91%) had heard of the COVID-19 vaccine (Table 2).

**Table 2.**
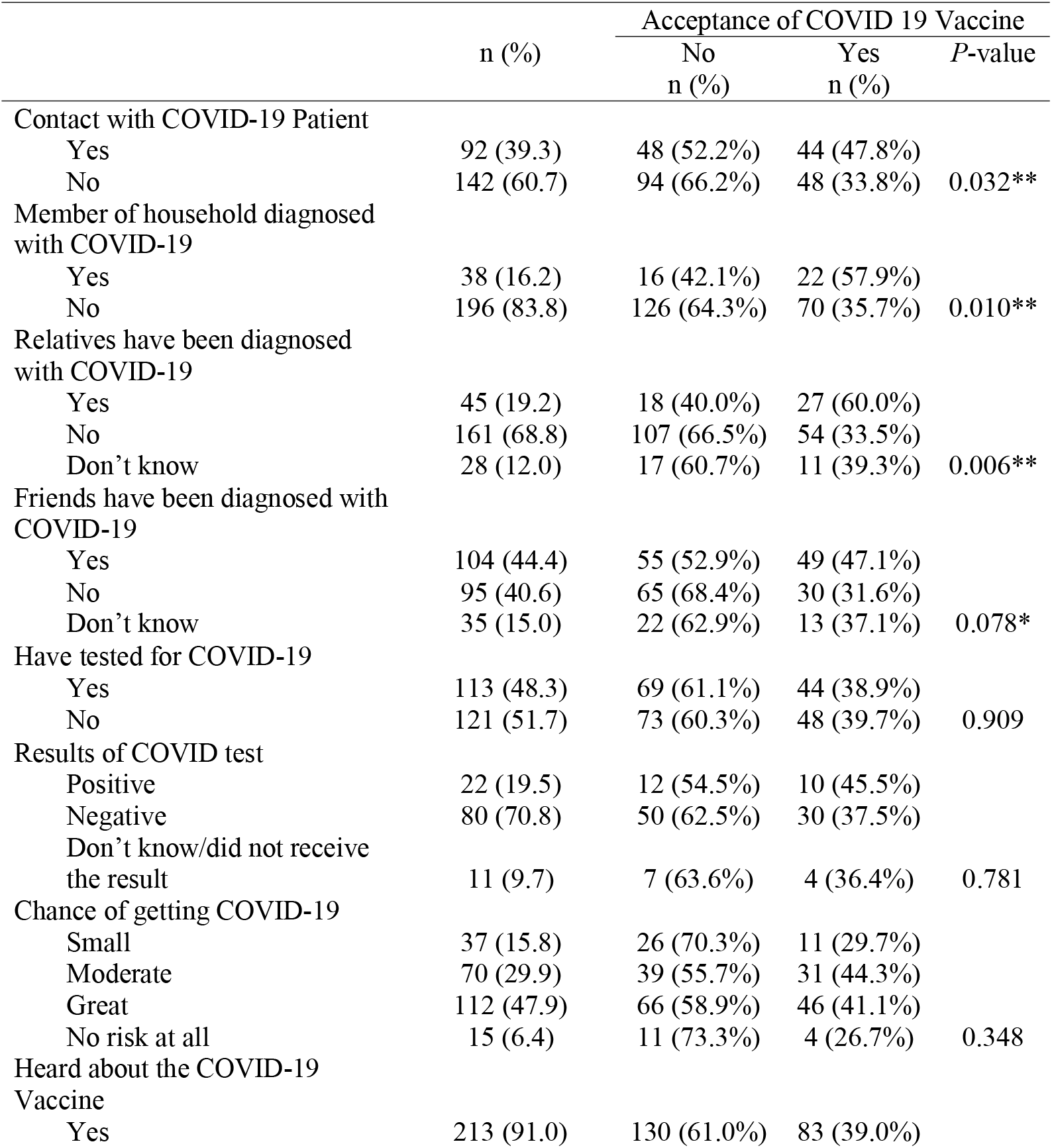
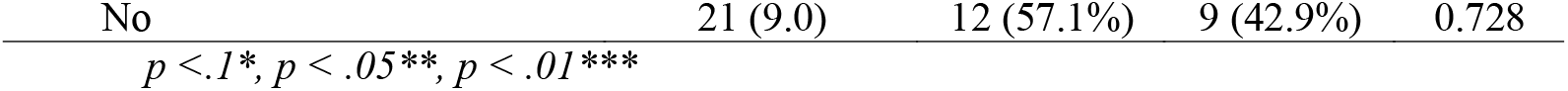
COVID-19 experience of health care workers

|  | n (%) | Acceptance of COVID 19 Vaccine |  |  |
| --- | --- | --- | --- | --- |
|  |  | No<br>n (%) | Yes<br>n (%) | *P*-value |
| Contact with COVID-19 Patient |  |  |  |  |
| Yes | 92 (39.3) | 48 (52.2%) | 44 (47.8%) | 0.032** |
| No | 142 (60.7) | 94 (66.2%) | 48 (33.8%) |  |
| Member of household diagnosed with COVID-19 |  |  |  |  |
| Yes | 38 (16.2) | 16 (42.1%) | 22 (57.9%) | 0.010** |
| No | 196 (83.8) | 126 (64.3%) | 70 (35.7%) |  |
| Relatives have been diagnosed with COVID-19 |  |  |  |  |
| Yes | 45 (19.2) | 18 (40.0%) | 27 (60.0%) | 0.006** |
| No | 161 (68.8) | 107 (66.5%) | 54 (33.5%) |  |
| Don't know | 28 (12.0) | 17 (60.7%) | 11 (39.3%) |  |
| Friends have been diagnosed with COVID-19 |  |  |  |  |
| Yes | 104 (44.4) | 55 (52.9%) | 49 (47.1%) | 0.078* |
| No | 95 (40.6) | 65 (68.4%) | 30 (31.6%) |  |
| Don't know | 35 (15.0) | 22 (62.9%) | 13 (37.1%) |  |
| Have tested for COVID-19 |  |  |  |  |
| Yes | 113 (48.3) | 69 (61.1%) | 44 (38.9%) | 0.909 |
| No | 121 (51.7) | 73 (60.3%) | 48 (39.7%) |  |
| Results of COVID test |  |  |  |  |
| Positive | 22 (19.5) | 12 (54.5%) | 10 (45.5%) | 0.781 |
| Negative | 80 (70.8) | 50 (62.5%) | 30 (37.5%) |  |
| Don't know/did not receive the result | 11 (9.7) | 7 (63.6%) | 4 (36.4%) |  |
| Chance of getting COVID-19 |  |  |  |  |
| Small | 37 (15.8) | 26 (70.3%) | 11 (29.7%) | 0.348 |
| Moderate | 70 (29.9) | 39 (55.7%) | 31 (44.3%) |  |
| Great | 112 (47.9) | 66 (58.9%) | 46 (41.1%) |  |
| No risk at all | 15 (6.4) | 11 (73.3%) | 4 (26.7%) |  |
| Heard about the COVID-19 Vaccine |  |  |  |  |
| Yes | 213 (91.0) | 130 (61.0%) | 83 (39.0%) |  |
| No | 21 (9.0) | 12 (57.1%) | 9 (42.9%) | 0.728 |
| $p < .1^*$, $p < .05^{**}$, $p < .01^{***}$ | | | | |

### 3.4 Social trust and acceptance of COVID-19 vaccine

The results of the study showed that about 42.3% of health care workers trusted the authorities in the fight against COVID-19, while 27.4% did not trust the authorities in the fight against COVID-19. Also, 58.1% of respondents had trust in the information provided by the media in the fight against COVID-19, while 18.8% of them had no trust in the information provided by the media in the fight against COVID-19. Also, about half (48.7%) of the health care workers had trust in the health care system and hospitals in the fight against COVID-19, while 32.6% indicated otherwise. Further, 45.7% of the health care workers had trust in the correct implementation of the measures to combat COVID-19, and about 39.3% trusted the appropriateness of the economic measures taken with COVID-19. A higher proportion (39.8%) of the health care workers disagreed with the statement that Ghana is more successful than the western countries in the fight against COVID-19 and more than half (56.4%) of health care workers disagreed that the COVID-19 pandemic will lose its effect with sunshine (Table 3).

**Table 3.** Social trust and acceptance of COVID-19 vaccine

|  | n (%) | Acceptance of COVID 19 Vaccine |  |  |
| --- | --- | --- | --- | --- |
|  |  | No<br>n (%) | Yes<br>n (%) | P-value |
| I trust in the authorities in the fight against COVID-19 |  |  |  |  |
| Disagree | 64 (27.4) | 40 (62.5%) | 24 (37.5%) | 0.703 |
| Neutral | 71 (30.3) | 45 (63.4%) | 26 (36.6%) |  |
| Agree | 99 (42.3) | 57 (57.6%) | 42 (42.4%) |  |
| I trust in the information provided by the media in the fight against COVID-19 |  |  |  |  |
| Disagree | 44 (18.8) | 28 (63.6%) | 16 (36.4%) | 0.449 |
| Neutral | 54 (23.1) | 36 (66.7%) | 18 (33.3%) |  |
| Agree | 136 (58.1) | 78 (57.4%) | 58 (42.6%) |  |
| I trust in our health system and hospitals in the fight against COVID 19 |  |  |  |  |
| Disagree | 65 (27.8) | 45 (69.2%) | 20 (30.8%) | 0.083* |
| Neutral | 55 (23.5) | 36 (65.4%) | 19 (34.6%) |  |
| Agree | 114 (48.7) | 61 (53.5%) | 53 (46.5%) |  |
| I trust in the accuracy of the measures taken by the government in the fight against COVID-19 |  |  |  |  |
| Disagree | 76 (32.6) | 51 (67.1%) | 25 (32.9%) | 0.041** |
| Neutral | 47 (20.0) | 33 (70.2%) | 14 (29.8%) |  |
| Agree | 111 (47.4) | 58 (52.2%) | 53 (47.8%) |  |
| I trust in the correct implementation of the measures in the fight against COVID-19 |  |  |  |  |
| Disagree | 72 (30.8) | 38 (52.8%) | 34 (47.2%) | 0.217 |
| Neutral | 55 (23.5) | 37 (67.3%) | 18 (32.7%) |  |
| Agree | 107 (45.7) | 67 (62.6%) | 40 (37.4%) |  |
| I trust in the appropriateness of the economic measures taken with respect to COVID-19 |  |  |  |  |
| Disagree | 71 (30.3) | 39 (54.9%) | 32 (45.1%) | 0.124 |
| Neutral | 71 (30.3) | 50 (70.4%) | 21 (29.6%) |  |
| Agree | 92 (39.4) | 53 (57.6%) | 39 (42.4%) |  |
| I think that we are more successful than the western countries in the fight against COVID-19 |  |  |  |  |
| Disagree | 93 (39.8) | 54 (58.1%) | 39 (41.9%) | 0.520 |
| Neutral | 63 (26.9) | 42 (66.7%) | 21 (33.3%) |  |
| Agree | 78 (33.3) | 46 (59.0%) | 32 (41.0%) |  |
| I think that sunshine will make COVID-19 lose its effect |  |  |  |  |
| Disagree | 132 (56.4) | 75 (56.8%) | 57 (43.2%) | 0.337 |
| Neutral | 62 (26.5) | 42 (67.7%) | 20 (32.3%) |  |
| Agree | 40 (17.1) | 25 (62.5%) | 15 (37.5%) |  |

$p < .1^*$ , $p < .05^{**}$ , $p < .01^{***}$

### 3.5 Factors associated with COVID-19 vaccine acceptance

Factors associated with COVID-19 vaccine acceptance among the health care workers are presented in Table 4. Only variables that were significant at the bivariate level were included in the model. The results showed that sex, category of health care workers, and marital status were significantly associated with acceptance of the COVID-19 vaccine (Model 1). Female health care workers were less likely to accept COVID-19 vaccine compare to their male counterparts (OR= 0.478; CI 95% 0.268 - 0.855; p = 0.013). Also, medical doctors were more likely to accept the COVID-19 vaccine than nurses and midwives (OR= 2.535; 95 CI%: 1.081 - 5.942; p = 0.032). Married health care workers were more likely to accept the COVID-19 vaccine than those who were not married (OR = 1.878; 95 CI%:1.073 - 3.280; p = 0.027).

**Table 4.**
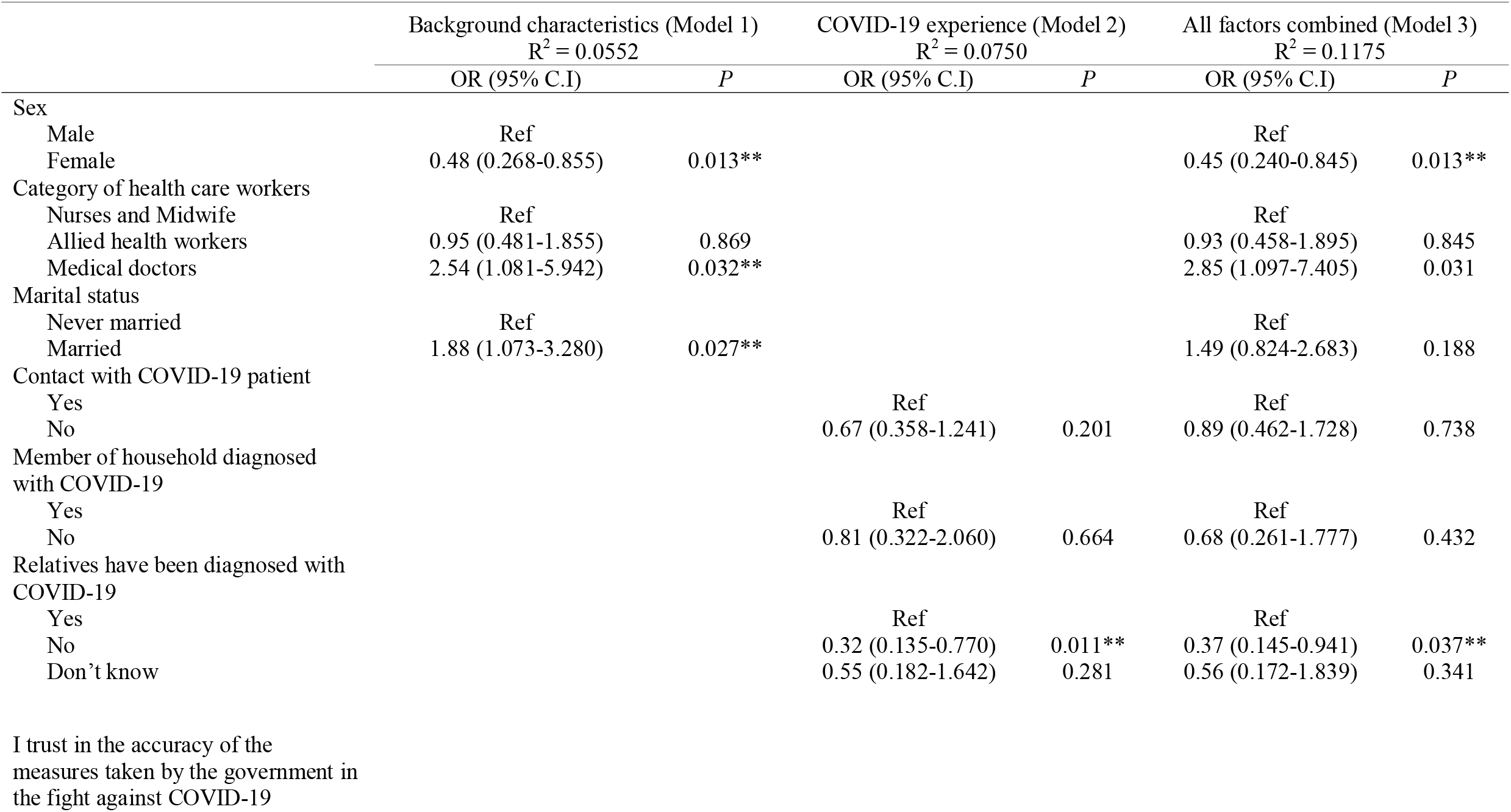
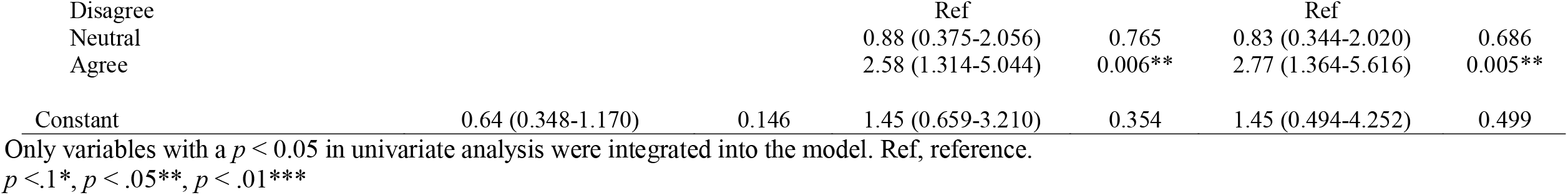
Logistic regression model for intention to accept COVID-19 vaccine

| | Background characteristics (Model 1)<br>$R^2 = 0.0552$ | | COVID-19 experience (Model 2)<br>$R^2 = 0.0750$ | | All factors combined (Model 3)<br>$R^2 = 0.1175$ | |
| --- | --- | --- | --- | --- | --- | --- |
|  | OR (95% C.I.) | *P* | OR (95% C.I.) | *P* | OR (95% C.I.) | *P* |
| Sex |  |  |  |  |  |  |
| Male | Ref |  |  |  | Ref |  |
| Female | 0.48 (0.268-0.855) | 0.013** |  |  | 0.45 (0.240-0.845) | 0.013** |
| Category of health care workers |  |  |  |  |  |  |
| Nurses and Midwife | Ref |  |  |  | Ref |  |
| Allied health workers | 0.95 (0.481-1.855) | 0.869 |  |  | 0.93 (0.458-1.895) | 0.845 |
| Medical doctors | 2.54 (1.081-5.942) | 0.032** |  |  | 2.85 (1.097-7.405) | 0.031 |
| Marital status |  |  |  |  |  |  |
| Never married | Ref |  |  |  | Ref |  |
| Married | 1.88 (1.073-3.280) | 0.027** |  |  | 1.49 (0.824-2.683) | 0.188 |
| Contact with COVID-19 patient |  |  |  |  |  |  |
| Yes |  |  | Ref |  | Ref |  |
| No |  |  | 0.67 (0.358-1.241) | 0.201 | 0.89 (0.462-1.728) | 0.738 |
| Member of household diagnosed with COVID-19 |  |  |  |  |  |  |
| Yes |  |  | Ref |  | Ref |  |
| No |  |  | 0.81 (0.322-2.060) | 0.664 | 0.68 (0.261-1.777) | 0.432 |
| Relatives have been diagnosed with COVID-19 |  |  |  |  |  |  |
| Yes |  |  | Ref |  | Ref |  |
| No |  |  | 0.32 (0.135-0.770) | 0.011** | 0.37 (0.145-0.941) | 0.037** |
| Don't know |  |  | 0.55 (0.182-1.642) | 0.281 | 0.56 (0.172-1.839) | 0.341 |
| I trust in the accuracy of the measures taken by the government in the fight against COVID-19 |  |  |  |  |  |  |
| Disagree |  |  | Ref |  | Ref |  |
| Neutral |  |  | 0.88 (0.375-2.056) | 0.765 | 0.83 (0.344-2.020) | 0.686 |
| Agree |  |  | 2.58 (1.314-5.044) | 0.006** | 2.77 (1.364-5.616) | 0.005** |
| Constant | 0.64 (0.348-1.170) | 0.146 | 1.45 (0.659-3.210) | 0.354 | 1.45 (0.494-4.252) | 0.499 |

Only variables with a $p < 0.05$ in univariate analysis were integrated into the model. Ref, reference.
$p < .1^*$ , $p < .05^{**}$ , $p < .01^{***}$

The results also showed that health care workers whose relative(s) had been diagnosed with COVID-19 and trust in the accuracy of the measures taken by the government in the fight against COVID-19 were significantly associated with acceptance of the COVID-19 vaccine (Model 2). Health care workers who indicated that they do not know if their relative(s) had been diagnosed with COVID-19 were less likely to accept the COVID-19 vaccine compared with health workers whose relative(s) had been diagnosed with COVID-19 (OR= 0.323; CI 95% 0.135 - 0.770; p = 0.011). Health care workers who agreed to the accuracy of measures taken by the government to fight COVID-19 were more likely to accept the COVID-19 vaccine than those who had no trust in the accuracy of the measures taken by the government in the fight against COVID-19 (OR= 2.575; CI 95%: 1.314 - 5.044; P = 0.006).

In Model 3, the adjusted results showed that sex, category of health care workers, relatives been diagnosed with COVID-19, and trust in the accuracy of measures taken by the government in the fight against COVID-19 were significantly associated with acceptance of the COVID-19 vaccine. Female health care workers were less likely to accept COVID-19 vaccine than males (OR= 0.451; CI 95% 0.240 - 0.845; p = 0.013). Also, medical doctors were more likely to accept the COVID-19 vaccine than nurses and midwife (OR=2.851; 95 CI%: 1.097 - 7.405; p = 0.031). Health care workers who indicated that they do not know if their relatives had been diagnosed with COVID-19 were less likely to accept the COVID-19 vaccine compared with those whose relatives had been diagnosed with COVID-19 (OR= 0.369; CI 95% 0.145 - 0.941; p = 0.037). Health care workers who agreed to the accuracy of measures taken by the government to fight COVID-19 were more likely to accept the COVID-19 vaccine than those who had no trust in the accuracy of the measures taken by the government in the fight against COVID-19 (OR= 2.768; CI 95%: 1.365 - 5.616; p = 0.005).

### 3.6 Reasons for non-acceptance of COVID-19 vaccines

The majority (64.5%) of health care workers were unwilling to accept the COVID-19 vaccine due to concern about the safety of the vaccines. Also, about 16% and 5% of them were unwilling to accept the COVID-19 vaccine due to concern after adverse effects of the vaccine, and acquiring COVID-19 through the vaccination, respectively (Table 5).

**Table 5.**
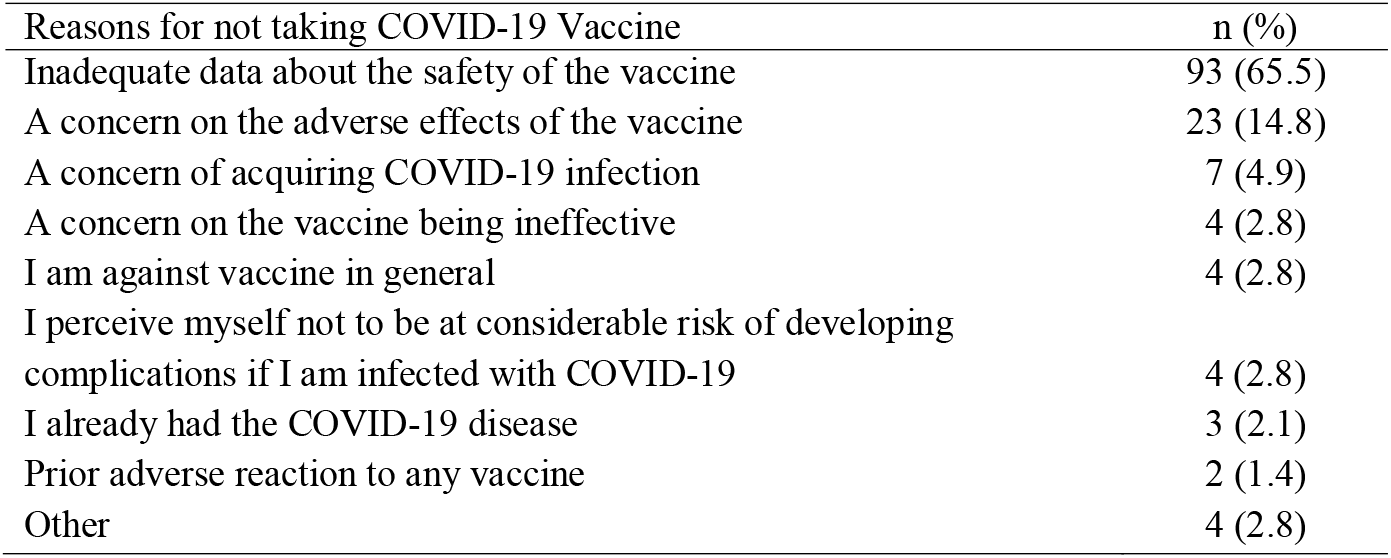
Reasons for non-acceptance of COVID-19 vaccines (N=142)

| Reasons for not taking COVID-19 Vaccine | n (%) |
| --- | --- |
| Inadequate data about the safety of the vaccine | 93 (65.5) |
| A concern on the adverse effects of the vaccine | 23 (14.8) |
| A concern of acquiring COVID-19 infection | 7 (4.9) |
| A concern on the vaccine being ineffective | 4 (2.8) |
| I am against vaccine in general | 4 (2.8) |
| I perceive myself not to be at considerable risk of developing complications if I am infected with COVID-19 | 4 (2.8) |
| I already had the COVID-19 disease | 3 (2.1) |
| Prior adverse reaction to any vaccine | 2 (1.4) |
| Other | 4 (2.8) |

## 4. Discussion

This study examined the acceptability of COVID-19 vaccines among health care workers in Ghana. The findings showed that approximately 39% of health care workers in Ghana intended to receive the COVID-19 vaccine if available. The acceptability of the COVID-19 vaccine in this study is comparable to that of Wang et al. [16] which found that about 40% of nurses in Hong Kong had the intention to accepting the COVID-19 vaccine if available. Similarly, in the multi-country study of Verger et al. [15], which also assessed health care workers’ attitudes towards COVID-19 vaccination in France, Belgium and Canada, it was found that approximately 40% of health care workers in Belgium (Wallonia and Brussels) were willing to vaccinate themselves if COVID-19 vaccines were available.

The acceptability of the COVID-19 vaccine in this study is higher compared to other countries in the SSA region. For example, the Nzaji et al. [17] study in the Democratic Republic of Congo found that approximately 28% of health care workers were willing to receive the COVID-19 vaccine if available. Conversely, the acceptability of the COVID-19 vaccine among health care workers in this study was lower than that reported in Saudi Arabia [25], China [26, 27], France [18], and the United States [13, 20, 28]. For instance, Shaw et al. [13] study in the United States found that about 58% of health care workers had the intention to receive the COVID-19 vaccine if available. The finding of low acceptance of the COVID-19 vaccine in the present study is worrying considering the associated consequences. Research has shown that health care workers intending to be vaccinated plan to recommend the vaccine to family and friends [19]. This suggests that low acceptance of the COVID-19 vaccine among health care workers may have a negative impact on the general population.

We also found that sex, category of health care workers, relative been diagnosed of COVID-19, and trust in the accuracy of the measures taken by the government in the fight against COVID-19 predicted acceptability of COVID-19 vaccination among health care workers in Ghana. The findings also revealed that female health care workers were less likely to accept COVID-19 vaccines if available compared to males. This finding corroborates other empirical studies that indicate that male health care workers are more likely to accept COVID-19 vaccines compared to female health care workers [13, 17-19]. The higher likelihood for male health care workers to accept COVID-19 vaccination has been attributed to increased risk perception of the disease in men compared to women [17]. A meta-analysis by Galbadage et al. [29] found that males disproportionately experience adverse outcomes and mortality from COVID-19 compared to females.

Medical doctors were more likely to accept COVID-19 vaccines if available compared to nurses and midwives. This finding is consistent with other studies that found that nurses are less likely to accept COVID-19 vaccines compared to doctors [13, 14, 17, 18]. According to Gagneux-Brunon et al. [18], the majority of nurses and midwives being females could explain why nurses and midwives are less likely to accept COVID-19 vaccines if available compared to medical doctors.

The findings also showed that health care workers whose relatives have not been diagnosed with COVID-19 were less likely to accept COVID-19 vaccines if available compared to health care workers whose relatives have been diagnosed with COVID-19. A probable explanation is that health care workers whose relatives have been diagnosed with COVID-19 may have gained more knowledge about COVID-19 and its effects on human health and consequently, they may want to protect themselves if COVID-19 vaccines become available.

Furthermore, the study found that health care workers who had trust in the accuracy of the measures taken by the government in the fight against COVID-19 were more likely to accept COVID-19 vaccines if available. During mass vaccination programmes, trust in authorities is key. The Strategic Advisory Group of Experts of the World Health Organization have identified trust in governments by citizens as essential for vaccine confidence as this helps reduce vaccine hesitancy [30]. The trust in the accuracy of other measures taken by the government of Ghana so far in the fight against COVID-19 disease may have influenced some health care workers to be willing to accept COVID-19 vaccines if they become available. Therefore, it is important for the government and health authorities to remain transparent to health care workers regarding measures in the fight against COVID-19 disease to strengthen trust between them and health care workers.

The study also revealed that concerns about the safety of vaccines, and adverse side effects of the vaccine were the main reasons why health care workers were unwilling to accept COVID-19 vaccines. These findings are consistent with other studies [13, 14, 19, 20]. For instance, a study in the Kingdom of Saud Arabia found concerns about the safety of vaccines and concern about side effects as the main reasons for unwillingness to accept COVID-19 vaccines [25]. The attitude and uptake of vaccination by health professionals is associated with the acceptance of vaccination by patients, compliance with vaccination schedules and thus reduces aversion or hesitation towards vaccination [17]. To enhance the acceptability of COVID-19 vaccination among health care workers and the general population in Ghana, policymakers in the health sector should address the concerns of health care workers about the safety and side effects of the COVID-19 vaccines as early as possible.

Also, interventions designed to address these concerns should be tailor-made taking into consideration the sex and category of health care workers.

## 5. Limitations

This study has some limitations. First, the study was cross-sectional and therefore causality cannot be established. Second, this study used convenient and snowball sampling techniques, which are non-probability sampling techniques, thus limiting the extent to which the results can be generalized to the population of health care workers in Ghana, although the online survey link was shared on health care workers’ WhatsApp platforms with members across the country. Notwithstanding these limitations, this study highlights the determinants of COVID-19 vaccine acceptability and concerns about the COVID-19 vaccine, which will help to reduce the hesitancy of the vaccine uptake among health care workers in Ghana.

## 6. Conclusion

This study demonstrates that interventions to promote vaccination of COVID-19 among health care workers in Ghana should take into account their socio-demographic characteristics (such as sex, and category of health care workers), COVID-19 experience, and trust in the measures taken by the government in the fight against COVID-19 to achieve the desired results.

## Data Availability

The data used in this study can be obtained on written request to the corresponding author.

## Data statement

The data used in this study can be obtained on written request to the corresponding author.

## Authorship statement

All authors certify that they meet the criteria for authorship of the ICMJE.

## Authors’ contributions

This study was conceived and designed by FK-A, MWA, and GFA-A. FK-A, MWA, GFA-A, and BA led the data collection. The data processing and analysis were conducted by MWA. The manuscript was prepared by FK-A, MWA, GFA-A and BA. All authors contributed to the editing and final approval of the manuscript for submission.

## Acknowledgement

The authors would like to thank all the health care workers who took time off their busy schedules to participate in the survey exercise.

